# Redesigning SARS-CoV-2 clinical RT-qPCR assays for wastewater RT-ddPCR

**DOI:** 10.1101/2021.03.02.21252754

**Authors:** Raul Gonzalez, Allison Larson, Hannah Thompson, Errin Carter, Xavier Fernandez Cassi

**Affiliations:** Hampton Roads Sanitation District, 1434 Air Rail Avenue, Virginia Beach, VA, 23455, United States; École Polytechnique Fédérale de Lausanne (EPFL), CH-1015 Lausanne, Switzerland

**Keywords:** SARS-CoV-2, Droplet Digital PCR, Wastewater, Wastewater-based Epidemiology, Wastewater Surveillance

## Abstract

COVID-19 wastewater surveillance has gained widespread acceptance to monitor community infection trends. Wastewater samples primarily differ from clinical samples by having low viral concentrations due to dilution, and high levels of PCR inhibitors. Therefore, wastewater samples should be processed by appropriately designed and optimized molecular workflows to accurately quantify targets. Digital PCR has shown to be more sensitive and resilient to environmental matrix inhibition. However, most SARS-CoV-2 assays have been designed for clinical use on RT-qPCR instruments, then adopted to digital PCR platforms. But it is unknown whether clinical RT-qPCR assays are adequate to use on digital PCR platforms. Here we designed an N and E gene multiplex (ddCoV_N and ddCoV_E) specifically for RT-ddPCR and benchmarked them against the nCoV_N2 and E_Sarbeco assays. ddCoV_N and ddCoV_E have equivalent limits of detections and wastewater sample concentrations to NCoV_N2 and E_Sarbeco but showed improved signal-to-noise ratios that eased interpretation and ability to multiplex. From GISAID downloaded unique sequences analyzed, 2.12% and 0.83% present a mismatch or would not be detected by the used primer/probe combination for the ddCoV_N and ddCoV_E, respectively.

## Introduction

Globally there have been over 83,000,000 cases and 1,800,000 deaths due to the coronavirus disease-2019 (COVID-19) pandemic in 2020 alone (Dong et al., 2020). Wastewater surveillance of severe acute respiratory syndrome coronavirus 2 (SARS-CoV-2), the virus that causes COVID-19, has been used as an epidemiological tool to better understand the infection extent in sewershed communities (e.g. Gerrity et al., 2021; Gonzalez et al., 2020; Prado et al., 2021; Randazzo et al., 2020; Hata et al. 2021; Carrillo-Reyes et al., 2020).

Two main differences exist between clinical and environmental samples. First, environmental samples typically have lower viral concentrations compared to clinical (e.g., nasopharyngeal, throat, stool, sputum) samples. This is primarily due to dilution effects and often require environmental samples to be concentrated prior to quantification. Second, environmental matrices can have a wide variety of inhibitory substances that are co-concentrated in environmental workflows. Thus, environmental samples require molecular workflows that have properly designed and optimized assays to maximize sensitivity and accuracy. An appropriate assay design reduces primer dimers that can interfere with accurate quantification, which is especially important at lower concentrations. Additionally, proper assay design and optimization can improve signal-to-noise ratios that can facilitate interpretation of true positives. The signal-to-noise ratio is especially important to certain digital PCR platforms (i.e., droplet digital PCR).

Reverse transcription-qPCR (RT-qPCR) and RT-digital PCR have been the dominant molecular methods used to quantify SARS-CoV-2 in wastewater (e.g., Pecson et al., 2021; Fores et al., 2021; Oliveira et al., 2020; Cervantes-Aviles et al., 2021; Alygizakis et al., 2021; Torii et al., 2021). Many RT-qPCR SARS-CoV-2 assays have been developed for various genome regions—N gene, E gene, S gene, etc. (Lu et al., 2020; Corman et al., 2020; Chu et al., 2020; China CDC, 2020). These assays have been designed based on general RT-qPCR specifications and some have design faults from not strictly following design guidelines. However, these assays can still provide adequate quantification, even without strict adherence to recommended design specifications, because of high titer clinical samples. Digital PCR has been proven to be more sensitive than qPCR (Falzone et al., 2020; Liu et al., 2020; Suo et al., 2020), especially in environmental matrices (Graham et al., 2021; Cao et al., 2015; Yang et al., 2014; Racki et al., 2014), with some exceptions (e.g., D’Aoust et al., 2021). The assays used have all been designed for clinical use on RT-qPCR instruments, then adopted to digital PCR platforms. Some digital PCR platforms, like droplet digital PCR (ddPCR), require a stricter adherence to assay design criterion since the technology relies on the clear separation of positive and negative droplets or partitions.

Early on in the SARS-CoV-2 pandemic the following knowledge gap has been identified: Are clinical RT-qPCR assays adequate to use on digital PCR platforms, especially when analyzing environmental samples that have strong matrix inhibition and are subject to dilution effects? Here we designed and optimized a SARS-CoV-2 RT-ddPCR multiplex for wastewater following strict design criteria to maximize accuracy especially at the lower concentrations in a difficult matrix (wastewater). N and E gene assays were specifically designed for ddPCR and compared to the nCoV_N2 and E_Sarbeco assays (Lu et al., 2020; Corman et al., 2020) designed for RT-qPCR. Knowing the effect of strict adherence to digital PCR design specifications is necessary since wastewater surveillance has seen a shift from trend analysis at the sewershed scale to finer resolution building level screening of individuals. Thus, pushing the need for an appropriately optimized method for environmental samples.

## Methods

### Assay Design

Many COVID-19 wastewater surveillance researchers have been running multiple assays for the same gene (e.g., Sherchan et al., 2020, Medema et al., 2020; Ahmed et al., 2020). This is highly redundant and not normally done in wastewater studies quantifying pathogens (e.g., Worley-Morse et al. 2019; Sarmila et al., 2020). A better strategy to gain higher confidence of results would be to quantify multiple conserved genes in the virus. We have designed 2 assays targeting N and E genes using Primer3Plus software (Untergasser et al., 2007) and the criteria found in Supplemental Table S1. In short, we strictly followed widely accepted PCR design criteria (and specific manufacturer recommendations) to lower primer dimer scores, and target recommended GC content (40-60%), melting temperatures (Tm; 60-63°C for primers, 4-6°C higher for probes), and lengths. The newly designed primers and probes for RT-ddPCR (ddCoV_N, ddCoV_E) can be seen in Table 1. Optimization and validation of these assays was done alongside the popularly used nCoV_N2 and E_Sarbeco assays. Geneious Prime (Geneious Biologics, Auckland, New Zealand) was used to visualize the newly designed primer and probe positions with respect to the nCoV_N2 and E_Sarbeco assays. Primer3Plus Primer_Check task (with the conditions in Supplementary Table S1) was used to determine the primer and probe Tm, GC content, and affinity for self-complementarity (likelihood that a primer will bind to itself or the other primer).

**Table 1.**
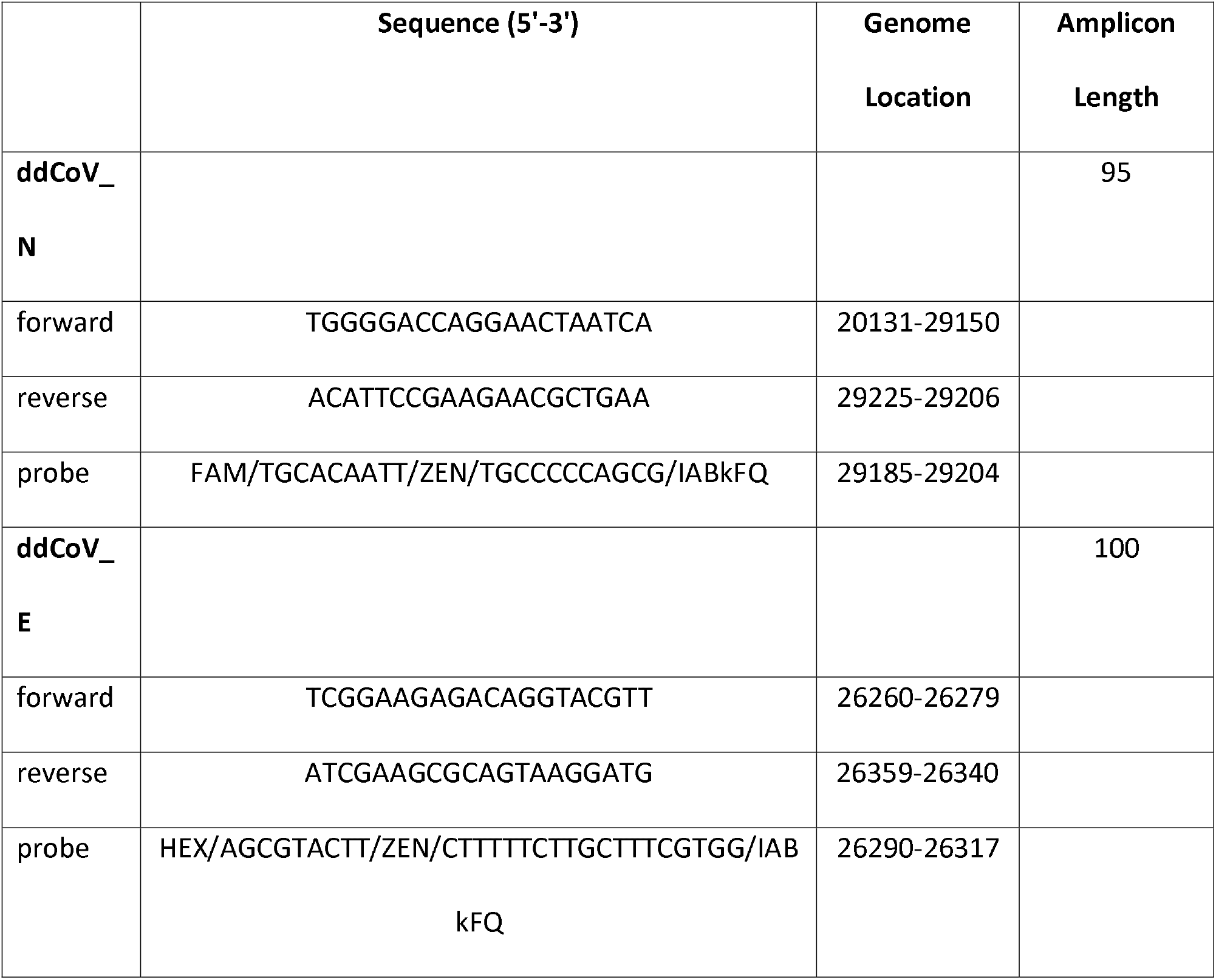
The N and E gene primer/probe sequences designed for RT-ddPCR. Genome locations based on SARS-CoV-2 (GenBank accession no. MN908947).

### Sample Concentration and RNA Extraction

Wastewater concentration was done using electronegative filtration according to Gonzalez et al. (2020). In short, MgCl_2_ was added to a final concentration of 25 mM to 25 - 50 mL wastewater, then acidified to a pH of 3.5 with 20% HCl. The sample was then filtered through a mixed cellulose ester HA filter (HAWP04700; Millipore, Billerica, MA, USA). A total of 1.0×10^6^ copies of bovine coronavirus (CALF-GUARD; Zoetis, Parsippany, NJ) was added to all samples prior to filtration to determine total process recovery. Bovine coronavirus recovery using RT-ddPCR (Gonzalez et al., 2020) was the ratio of bovine coronavirus sample concentration to spike concentration.

Filters were immediately stored in a -80°C freezer until RNA extraction on a NucliSENS easyMag (bioMerieux, Inc., Durham, NC, USA) within 5 days of filtration. Extractions were preformed using a modified manufacturer’s protocol B 2.0.1. Modifications included a 30-min off board lysis (2 mL of lysis buffer) and 100 μL of magnetic silica beads to minimize matrix inhibition and maximize RNA recovery.

### Reverse Transcription Droplet Digital PCR

Digital PCR workflow used the one-step RT-ddPCR advanced kit for probes on the Bio-Rad QX200 (Bio-Rad, Hercules, CA, USA). The 20 μL final reaction volume consisted of the following: 5 μL 1 × one-step RT-ddPCR Supermix (Bio-Rad), 2 μL reverse transcriptase (Bio-Rad), 1 μL 300 mM DTT, 3 μL of forward and reverse primers (900 nM final concentration) and probes (250 nM final concentration), 5 μL RNase-free water, and 4 μL RNA (samples run undiluted). The 20 uL volume was combined with 70 μL droplet generation oil in the Droplet Generator (Bio-Rad) and resulting droplets were transferred to a 96-well plate for end-point PCR. PCR cycling conditions were: 60-min reverse transcription at 50°C (1 cycle), 10-min enzyme activation at 95°C (1 cycle), 30-s denaturation at 94°C (40 cycles), 1-min annealing/extension cycle at 55°C (40 cycles; ramp rate of ~ 2–3°C/s), 10-min enzyme deactivation at 98°C (1 cycle) and a hold at 4°C until read on the droplet reader. Positive and negative droplet reading occurred on the Bio-Rad Droplet Reader.

### Thermal Gradient Optimization

Optimizing the annealing temperature of a PCR assay is critical, especially for target specificity. The C1000 Thermal Cycler (Bio-Rad) was used to test the reaction across a range of temperatures (60 to 50°C). The Twist Synthetic SARS-CoV-2 RNA Control 4 (part no. 102862; Twist Bioscience, San Francisco, CA, USA) and a SARS-CoV-2 positive wastewater sample was used for the optimization. This optimization also gives the first visualization at the assay efficiency and robustness.

### Limits of Detection

Theoretical limits of detection (LOD) were calculated by running serial dilutions of the Twist Synthetic SARS-CoV-2 RNA Control 4 (Twist Bioscience, San Francisco, CA, USA) standard in 10 replicates over 4 orders of magnitude. The LOD was the concentration (copies/reaction) at which over 60% of the technical replicates were positive (Gonzalez et al., 2020). Coefficient of variations (CV) were calculated for the copies per reaction for the serial dilutions to compare replicates at different positive droplet concentrations.

### In-silico specificity and primer/probe mismatch analysis

BLASTn (Altschul et al., 1990) was used to test the in-silico specificity of the ddCoV_N and ddCoV_E primers against viruses (taxid: 10239) and bacteria (taxid: 2) in the nr database. There were no BLAST hits presenting good homology and coverage for the primer/probe combinations that suggests assay detection to viruses or bacteria other than SARS-CoV-2.

A total of 105,268 SARS-CoV-2 sequences retrieved and downloaded from GISAID on 06/02/2021 were evaluated in silico for the primer/probes designed. The inclusion criteria for these sequences to be tested in silico were: 1) uploaded sequences from the United States of America on date 01/01/2020 to 31/12/2020; 2) worldwide uploaded sequences from 01/01/2021 to 06/02/2021; 3) only those who have been sequenced from a human host and the collection date is included; 5) only fully completed SARS-CoV-2 genomes from all clades and lineages presenting high coverage were included, low coverage genomes were excluded. Briefly, sequences were imported in Geneious Prime (Geneious Biologics) and duplicated sequences with identical names were removed. Using Geneious software, newly designed primers/probes for the nucleocapsid protein (ddCoV_N) and the envelope gene (ddCoV_E) were mapped using Geneious primer mapper against the retrieved sequences allowing a maximum of 3 mismatches. Sequences with more than three mismatches are summarized in Supplemental Figure S1 and could be considered unlikely to be detected by the tested assay. A list of complete mismatches and the relative position within SARS-CoV-2 genome are presented in Supplemental Table S2.

### Concentration Comparison

Concentration comparison of the two new assays, nCoV_N2, and E_Sarbeco assays was done on 3 weeks of wastewater samples from 9 different facilities (N=27). These facilities and the collection scheme are described in Gonzalez et al. (2020). Within gene assays concentrations were compared using percent difference.

## Results and Discussion

### Assays and Performance

The newly designed primers and probes (ddCoV_N and ddCoV_E) are adjacent to the respective nCoV_N2 and E_Sarbeco assay locations. Figure 1 documents the shift in primers and probes necessary to strictly meet widely accepted PCR design criteria and specific manufacturer recommendations (Supplemental Table S1). The information for the two newly designed RT-ddPCR primers and probes as well as the nCoV-N2 and E_Sarbeco assays are in Table 2. The nCoV_N2 and E_Sarbeco assays have high self-complementarity scores in the forward primers. The newly designed assays reduced this score as well as restricted within assay primer/probe Tm differences and constricted the GC%.

**Table 2.** Primer and probe sequence information. Sequences with an asterisk (*) exhibits high end self complementarity.

|  | Tm (°C) | GC (%) | ANY | SELF |
| --- | --- | --- | --- | --- |
| **ddCoV_N** |  |  |  |  |
| probe | 66.8 | 60 | 6 | 2 |
| forward | 60 | 50 | 3 | 2 |
| reverse | 60.1 | 45 | 5 | 0 |
| **ddCoV_E** |  |  |  |  |
| probe | 66.9 | 42.9 | 4 | 0 |
| forward | 60.4 | 50 | 4 | 2 |
| reverse | 60.4 | 50 | 4 | 0 |
| **nCoV_N2** |  |  |  |  |
| probe | 68.8 | 56.5 | 6 | 2 |
| forward* | 59.4 | 40 | 5 | 5 |
| reverse | 60.7 | 55.6 | 5 | 2 |
| **E_Sarbeco** |  |  |  |  |
| probe | 69.1 | 53.8 | 4 | 2 |
| forward* | 60.6 | 34.6 | 8 | 8 |
| reverse | 63.2 | 45.5 | 7 | 1 |

**Figure 1.**
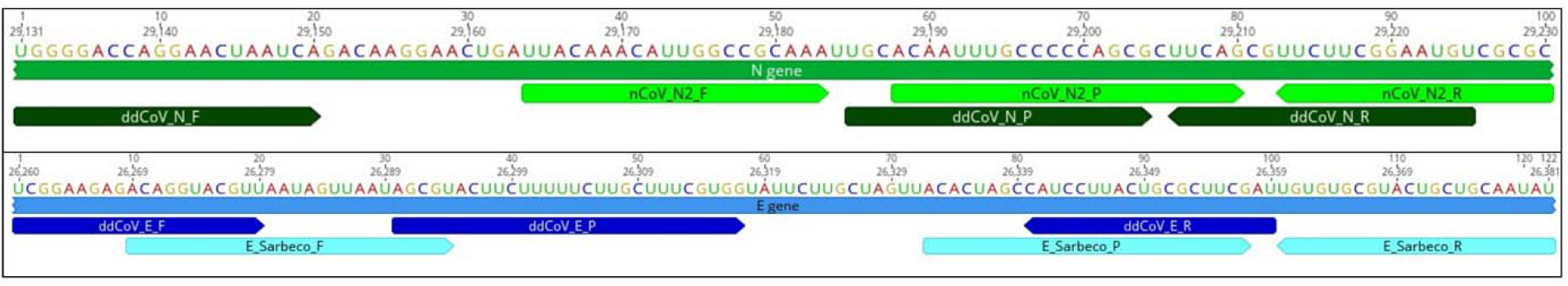
Locations of RT-ddPCR and RT-qPCR designed assays with respect to the SARS-CoV-2 genome (GenBank accession no. MN908947). ddCoV_N and nCoV_N2 assays are located on the N gene (top panel). ddCoV_E and E_Sarbeco are located on the E gene (bottom panel).

The annealing temperature thermal gradients give the first look at the efficiencies of the 4 assays on the QX200 ddPCR platform (Figure 2). With respect to the N gene assays, ddCoV_N created a superior separation between positive and negative droplets for both the high titer standard and wastewater sample. Positive droplet amplitude was approximately 10000 for both assays, while the negative droplet band was approximately 500 and 6500 for ddCoV_N and nCoV_N2, respectively. Additionally, the baseline for the ddCoV_N assay was significantly tighter. During the annealing temperature optimization, droplets in the ddCoV_N negative amplitude ranged from approximately 0 to 1000. The droplets in the nCoV_N2 ranged from approximately 500 to 7000. In the wastewater samples, ‘raininess’ was diminished with the ddCoV_N assay. During the optimization, the E gene assays did not have as significant difference in band amplitudes. The E_Sarbecco amplitude difference was slightly greater than that of the ddCoV_E. E_Sarbeco ranged from approximately 6500 to 11500, while the ddCoV_E ranged from approximately 1600 to 4600. However, the ddCoV_E gene had a noticeably tighter negative baseline—approximately half of the E_Sarbeco assay.

**Figure 2.**
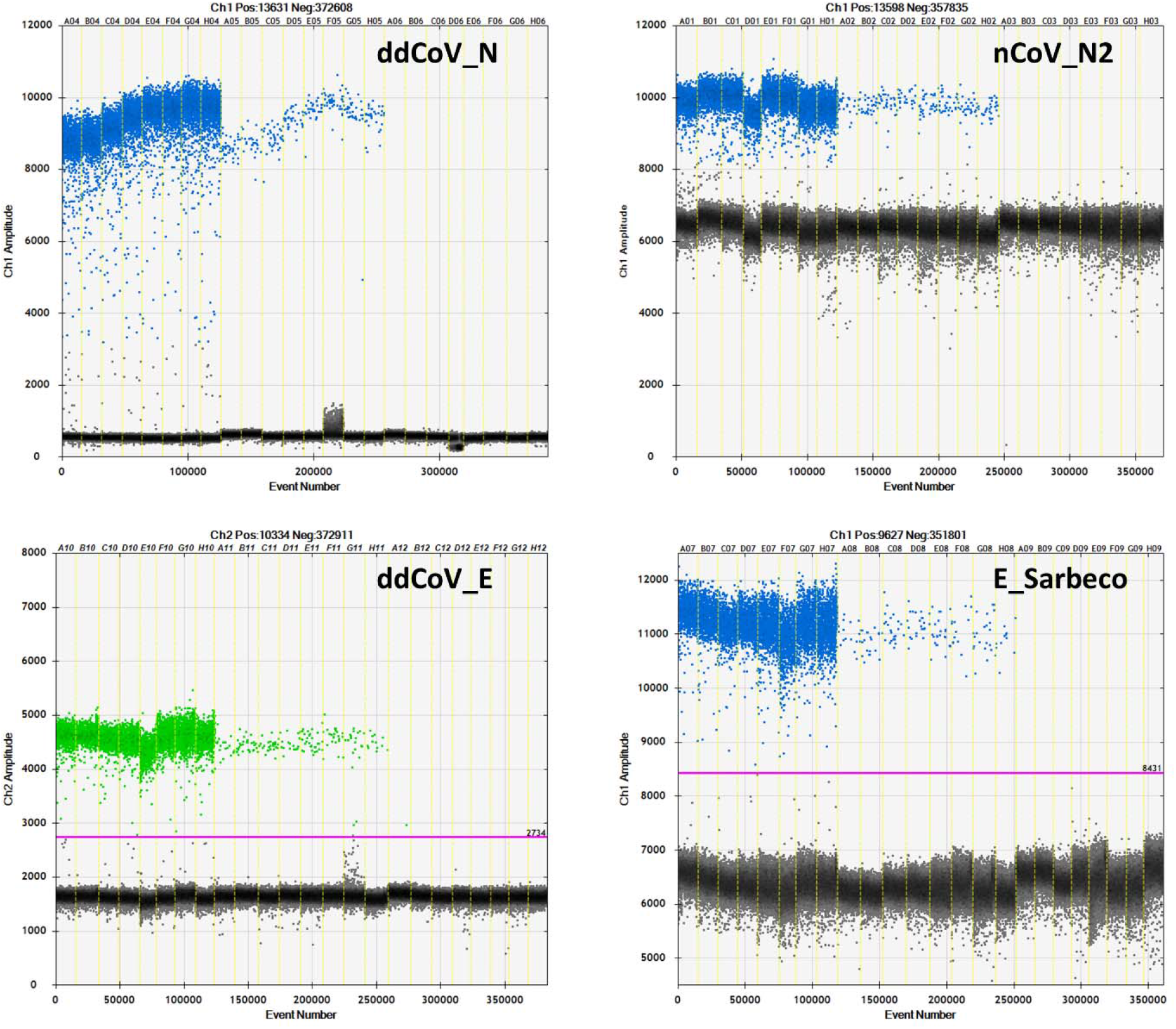
Annealing temperature thermal gradient optimizations for the 2 RT-ddPCR designed assays (ddCoV_N, ddCoV_E) and the 2 RT-qPCR designed assays (nCoV_N2, E_Sarbeco). Thicker portion of the positive band (first 8 wells) used RNA oligo standard, while the thinner portion (next 8 wells) used a SARS-CoV-2 positive wastewater sample. The last 8 wells are negatives (NTC).

The increased droplet band separation of the ddCoV_N assay and the tighter negative bands allows for greater ease in interpretation of results. Manual placement of the threshold is difficult with a small gap between positive and negative bands. This can be exacerbated with the ‘raininess’ caused by partial fluorescence and loose bands. These favorable characteristics are also essential to multiplexing assays. The increased separation and reduction in ‘raininess’ are likely in part due to the elimination of the BHQ (LGC Biosearch Technologies, Risskov, Denmark) quencher. While recommended by the manufacturer for the instrument, Bio-Rad has advised against the use of BHQ probes (LGC Biosearch Technologies) with their one-step reverse transcription kits since it contains Dithiothreitol (DTT). DTT is thought to cleave BHQ (LGC Biosearch Technologies) when not intended, causing fluorescence clusters. It is unlikely that the ZEN/Iowa Black FQ (Integrated DNA Technologies, Coralville, Iowa) double quenched probes alone caused the tighter baselines since the BHQ (LGC Biosearch Technologies) probes used for the nCoV-N2 and E_Sarbeco probes were also double quenched BHQnova (LGC Biosearch) probes.

### Limits of Detection

The LODs were nearly identical for each pair of gene assays but almost an order of magnitude different across genes. The N gene LODs were 2.16 and 2.14 copies per reaction for the ddCoV_N and nCoV_N2 assays, respectively. The E gene LODs were 16.0 and 15.0 copies per reaction for the ddCoV_E and E_Sarbeco assays, respectively. Figure 3 shows ddCoV_N and ddCoV_E number of positive droplets and the copies per reaction for the serial dilutions used to calculate the LODs. Coefficients of variation for the replicates were computed using the copies per reaction data for each dilution. For both assays the CV increased through the dilution series but took a large increase at the LODs. The CV increased faster in the ddCoV_E, than the ddCoV_N assay, reflecting the higher LOD.

**Figure 3.**
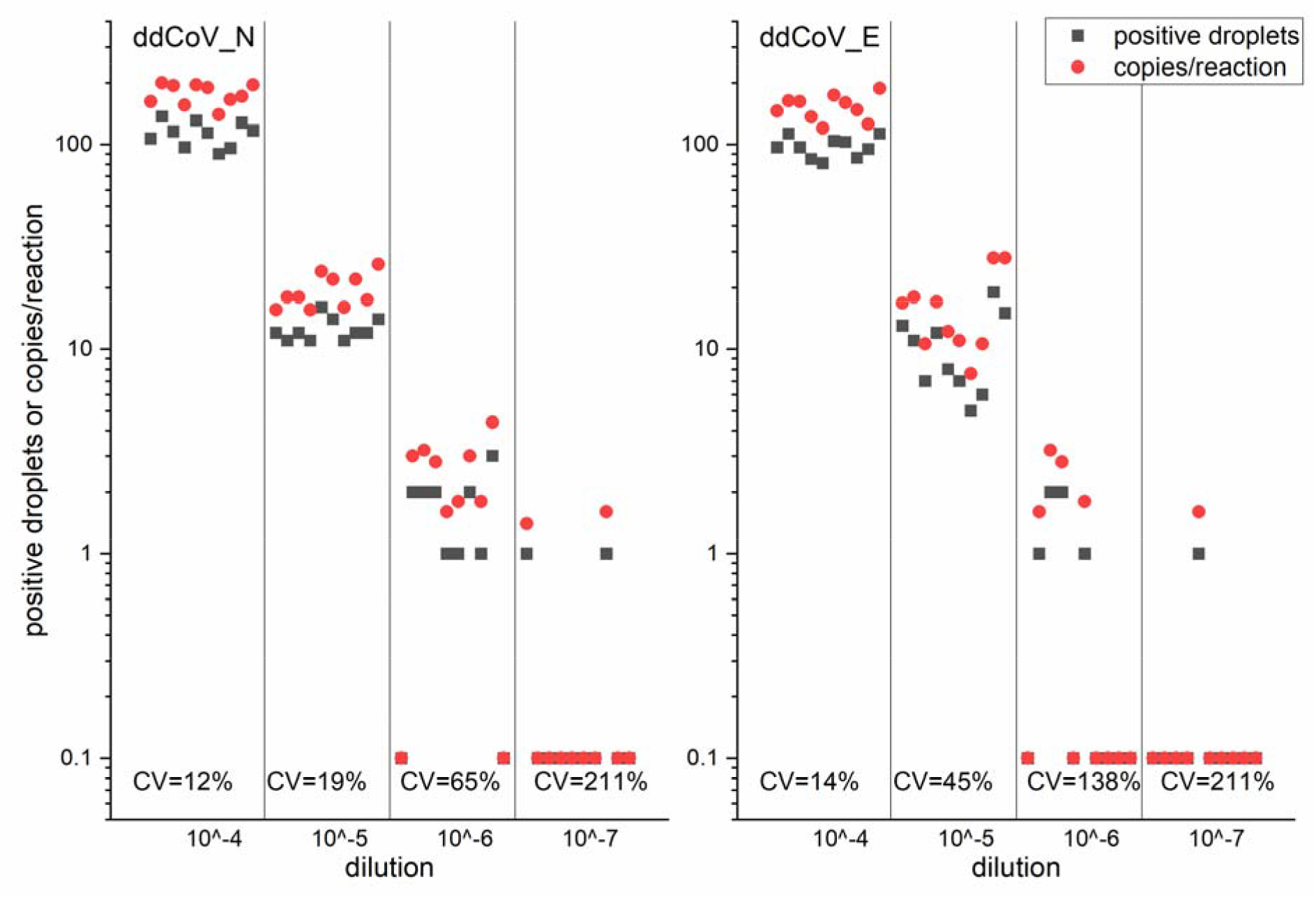
ddCoV_N and ddCoV_E positive droplets and copies per reaction for the limits of detection serial dilutions. Coefficients of variation (CV) for the copies per reaction are shown.

### In-Silico Analyses

From the total of unique sequences analysed, 869 out of 105,227 (0.83%) present a mismatch or would not be detected by the used primer/probe combination for the E gene (ddCoV_E). For E gene assay, most of the mutations affected the designed probe (55%) and the forward primer (35%).

For the in-silico testing of ddCoV_N assay, the total number of sequences analysed was reduced as the retrieved GISAID sequences presented ambiguous basecalls at the Nucleocapsid region. This was not observed for the E gene and it is likely that these ambiguities are related to a high mutation rate on this specific genome site or the fact that the N gene is located at the 3’ of SARS-CoV-2 genome. These ambiguities produced false positive results on the primer/probe mapping, significantly increasing the percentage of mismatches detected. Therefore, from the initial 105,227 sequences, a trimming on the sequences on the 3’ was applied when more than 40 continuous ambiguous base call were found. As a result of this, some sequences were significantly shortened and only genomes longer than 28Kbs bases were included in the in-silico test for the ddCoV_N. This reduced the total initial sequences analysed to 89,566. From the total of sequences included, 1901 (2.12%) presented a mismatch or would not be detected by the designed assay (more than 3 mismatches). Most of the identified mismatches (1901) affected the probe (47%) or the primer forward (41.6%) targeting the N-gene.

### Concentration Comparison

For three weeks the wastewater of 9 major facilities was analyzed using all 4 assays (Figure 4). Bovine coronavirus recoveries for the samples ranged from 0.38% to 19%, with a mean (SD) of 6.6% (4%). These recoveries are in line with previously reported recoveries for the same virus and workflow used (Gonzalez et al., 2020). The detectable concentrations across all assays ranged from 7.00×10^1^ to 2.21×10^4^ copies/100 mL. One sample was non-detect for all assays. Specifically, the mean (SD) detections for ddCoV_N and nCoV_N2 assays were 4.67×10^3^ (3.60×10^3^) and 5.46 ×10^3^ (4.42 ×10^3^), respectively. The mean (SD) detections for ddCoV_E and E_Sarbeco assays were 1.94 ×10^3^ (1.46 ×10^3^) and 1.92 ×10^3^ (1.49 ×10^3^), respectively. The mean (SD) percent difference for detections (N=26) using the N assays was 12 (19). The mean percent difference between the E gene assays was 7 (32). Differences within gene assays was small as expected since the assays were designed around the same genomic locations. The 2-D plot (Figure 5) highlights the assays multiplexing performance for the first 9 samples analyzed. There is clear clustering of the droplet populations—gray = double negative, blue = FAM positive, green = HEX positive, and orange = double positive. The optimized, newly designed assays translate well to multiplexing and easily interpretable results.

**Figure 4.**
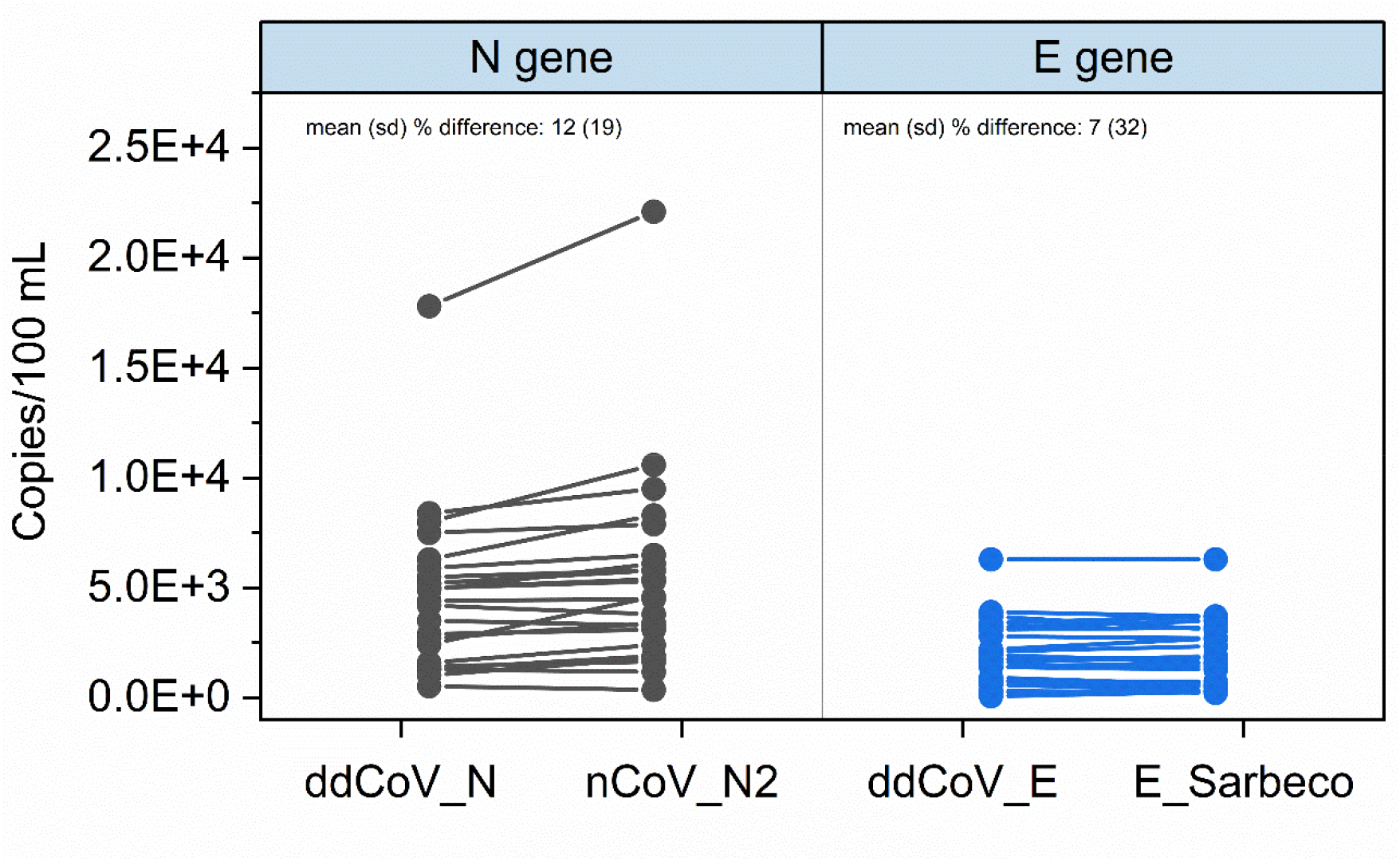
Assay comparisons for three weeks of wastewater SARS-CoV-2 concentrations at 9 facilities in southeast Virginia. Only the detectable concentrations are shown (N=26).

**Figure 5.**
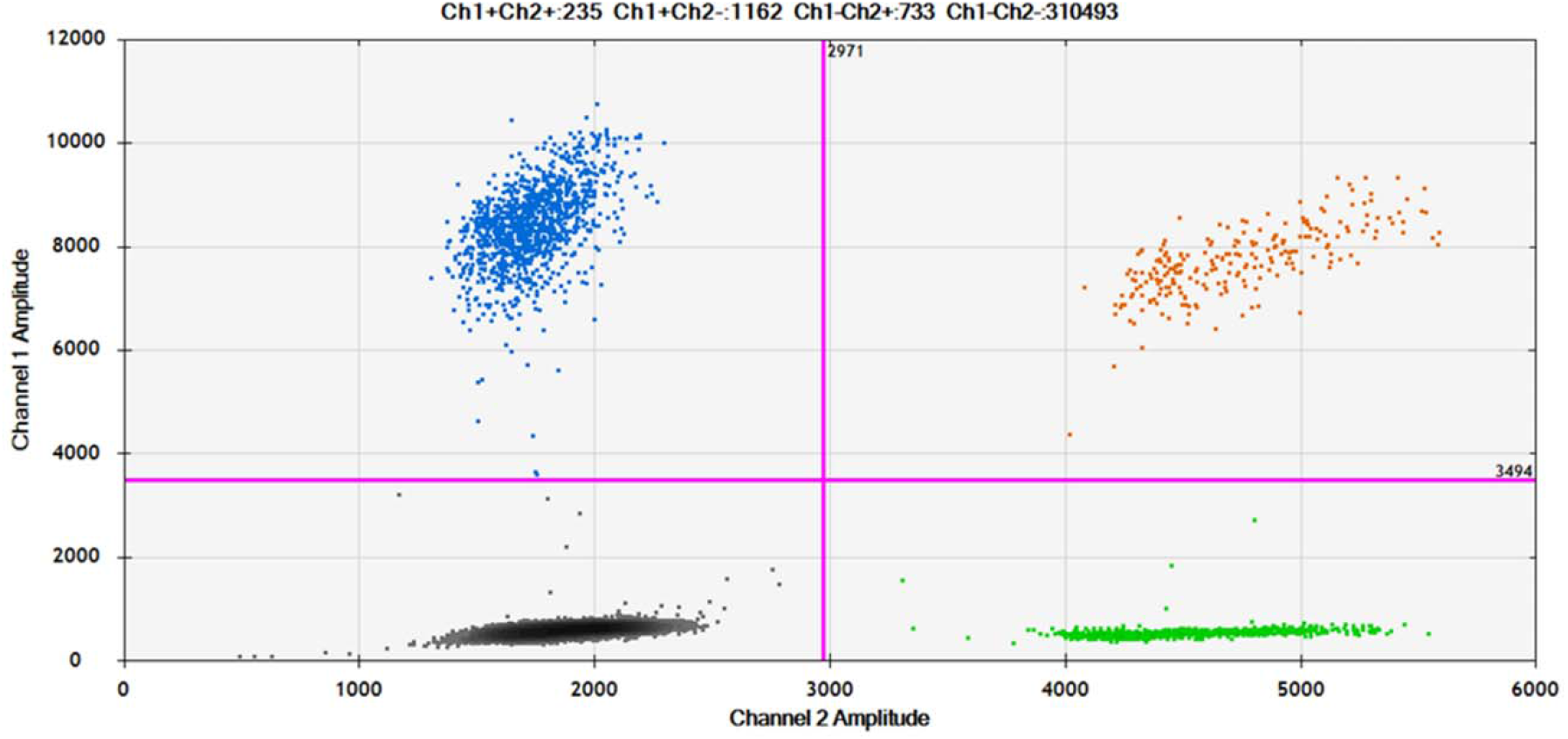
The 2-D plot of the ddCoV_N and ddCoV_E multiplexed assays for the first 9 samples analyzed. Efficient clustering of the droplet populations is shown—gray = double negative, blue = FAM positive, green = HEX positive, and orange = double positive.

## Conclusion

SARS-CoV-2 wastewater surveillance has been practiced widely to examine community level infection trends and even to screen for infected individuals at the building level. Wastewater SARS-CoV-2 concentrations can be significantly lower than clinical samples due to dilution and environmental RNA degradation. Because of this well designed and optimized molecular assays should be used in environmental sample workflows. The ddCoV_N and ddCoV_E assays have been designed to lower dimer scores and strictly follow PCR design guidelines, specifically those recommended for ddPCR. While the RT-ddPCR assays have equivalent LOD and concentrations, they have improved signal-to-noise ratios that ease interpretation and decrease the likelihood of false positives due to misinterpretation. This is especially important for novice users and ultimately this improved efficiency is translated to an easier ability to multiplex. *In-silico* analysis of the designed primers and probes should be periodically conducted to ensure continued assay appropriateness in SARS-CoV-2 environmental monitoring. New variants are constantly emerging with mutations affecting the targeted genes which ultimately can decrease the sensitivity of currently used assays.

## Data Availability

Raw data is available upon request.

## Declarations

### Availability of data and material

This manuscript has data included as supplemental information. Additional data will be made available on reasonable request.

## References

Ahmed, W., Angel, N., Edson, J., Bibby, K., Bivins, A., O’Brien, J.W., Choi, P.M., Kitajima, M., Simpson, S.L., Li, J. and Tscharke, B., 2020. First confirmed detection of SARS-CoV-2 in untreated wastewater in Australia: a proof of concept for the wastewater surveillance of COVID-19 in the community. Science of the Total Environment, 728, p.138764.

Altschul, S.F., Gish, W., Miller, W., Myers, E.W. and Lipman, D.J., 1990. Basic local alignment search tool. Journal of molecular biology, 215(3), pp.403–410.

Alygizakis, N., Markou, A.N., Rousis, N.I., Galani, A., Avgeris, M., Adamopoulos, P.G., Scorilas, A., Lianidou, E.S., Paraskevis, D., Tsiodras, S. and Tsakris, A., 2020. Analytical methodologies for the detection of SARS-CoV-2 in wastewater: Protocols and future perspectives. TrAC Trends in Analytical Chemistry, p.116125.

Cao, Y., Raith, M.R. and Griffith, J.F., 2015. Droplet digital PCR for simultaneous quantification of general and human-associated fecal indicators for water quality assessment. water research, 70, pp.337–349.

Carrillo-Reyes, J., Barragán-Trinidad, M. and Buitrón, G., 2020. Surveillance of SARS-CoV-2 in sewage and wastewater treatment plants in Mexico. Journal of Water Process Engineering, p.101815.

Cervantes-Avilés, P., Moreno-Andrade, I. and Carrillo-Reyes, J., 2021. Approaches applied to detect SARS-CoV-2 in wastewater and perspectives post-COVID-19. Journal of Water Process Engineering, p.101947.

Chu, D.K., Pan, Y., Cheng, S.M., Hui, K.P., Krishnan, P., Liu, Y., Ng, D.Y., Wan, C.K., Yang, P., Wang, Q. and Peiris, M., 2020. Molecular diagnosis of a novel coronavirus (2019-nCoV) causing an outbreak of pneumonia. Clinical chemistry, 66(4), pp.549–555.

Corman, V.M., Landt, O., Kaiser, M., Molenkamp, R., Meijer, A., Chu, D.K., Bleicker, T., Brünink, S., Schneider, J., Schmidt, M.L. and Mulders, D.G., 2020. Detection of 2019 novel coronavirus (2019-nCoV) by real-time RT-PCR. Eurosurveillance, 25(3), p.2000045.

D’Aoust, P.M., Mercier, E., Montpetit, D., Jia, J.J., Alexandrov, I., Neault, N., Baig, A.T., Mayne, J., Zhang, X., Alain, T. and Langlois, M.A., 2021. Quantitative analysis of SARS-CoV-2 RNA from wastewater solids in communities with low COVID-19 incidence and prevalence. Water research, 188, p.116560.

Dong, E., Du, H. and Gardner, L., 2020. An interactive web-based dashboard to track COVID-19 in real time. The Lancet infectious diseases, 20(5), pp.533–534.

Falzone, L., Musso, N., Gattuso, G., Bongiorno, D., Palermo, C.I., Scalia, G., Libra, M. and Stefani, S., 2020. Sensitivity assessment of droplet digital PCR for SARS-CoV-2 detection. International journal of molecular medicine, 46(3), pp.957–964.

Forés, E., Bofill-Mas, S., Itarte, M., Martínez-Puchol, S., Hundesa, A., Calvo, M., Borrego, C.M., Corominas, L.L., Girones, R. and Rusiñol, M., 2021. Evaluation of two rapid ultrafiltration-based methods for SARS-CoV-2 concentration from wastewater. Science of The Total Environment, 768, p.144786.

Gerrity, D., Papp, K., Stoker, M., Sims, A. and Frehner, W., 2021. Early-pandemic wastewater surveillance of SARS-CoV-2 in Southern Nevada: Methodology, occurrence, and incidence/prevalence considerations. Water research X, 10, p.100086.

Gonzalez, R., Curtis, K., Bivins, A., Bibby, K., Weir, M.H., Yetka, K., Thompson, H., Keeling, D., Mitchell, J. and Gonzalez, D., 2020. COVID-19 surveillance in Southeastern Virginia using wastewater-based epidemiology. Water research, 186, p.116296.

Graham, K.E., Loeb, S.K., Wolfe, M.K., Catoe, D., Sinnott-Armstrong, N., Kim, S., Yamahara, K.M., Sassoubre, L.M., Mendoza Grijalva, L.M., Roldan-Hernandez, L. and Langenfeld, K., 2020. SARS-CoV-2 RNA in Wastewater Settled Solids Is Associated with COVID-19 Cases in a Large Urban Sewershed. Environmental science & technology.

Hata, A., Hara-Yamamura, H., Meuchi, Y., Imai, S. and Honda, R., 2021. Detection of SARS-CoV-2 in wastewater in Japan during a COVID-19 outbreak. Science of The Total Environment, 758, p.143578.

Liu, X., Feng, J., Zhang, Q., Guo, D., Zhang, L., Suo, T., Hu, W., Guo, M., Wang, X., Huang, Z. and Xiong, Y., 2020. Analytical comparisons of SARS-COV-2 detection by qRT-PCR and ddPCR with multiple primer/probe sets. Emerging microbes & infections, 9(1), pp.1175–1179.

Lu, X., Wang, L., Sakthivel, S.K., Whitaker, B., Murray, J., Kamili, S., Lynch, B., Malapati, L., Burke, S.A., Harcourt, J. and Tamin, A., 2020. US CDC real-time reverse transcription PCR panel for detection of severe acute respiratory syndrome coronavirus 2. Emerging infectious diseases, 26(8), p.1654.

Medema, G., Heijnen, L., Elsinga, G., Italiaander, R. and Brouwer, A., 2020. Presence of SARS-Coronavirus-2 RNA in sewage and correlation with reported COVID-19 prevalence in the early stage of the epidemic in the Netherlands. Environmental Science & Technology Letters, 7(7), pp.511–516.

National Institute for Viral Disease Control and Prevention. Specific primers and probes for detection of 2019 novel coronavirus (2020); http://ivdc.chinacdc.cn/kyjz/202001/t20200121_211337.html

Oliveira, M.D.L.A., Campos, A., Matos, A.R., Rigotto, C., Martins, A.S., Teixeira, P.F. and Siqueira, M.M., 2020. Wastewater-Based Epidemiology (WBE) and Viral Detection in Polluted Surface Water: A Valuable Tool for COVID-19 Surveillance—A Brief Review.

Pecson, B.M., Darby, E., Haas, C., Amha, Y., Bartolo, M., Danielson, R., Dearborn, Y., Di Giovanni, G., Ferguson, C., Fevig, S. and Gaddis, E., 2021. Reproducibility and sensitivity of 36 methods to quantify the SARS-CoV-2 genetic signal in raw wastewater: findings from an interlaboratory methods evaluation in the US. Environmental Science: Water Research & Technology.

Prado, T., Fumian, T.M., Mannarino, C.F., Resende, P.C., Motta, F.C., Eppinghaus, A.L.F., do Vale, V.H.C., Braz, R.M.S., de Andrade, J.D.S.R., Maranhão, A.G. and Miagostovich, M.P., 2021. Wastewater-based epidemiology as a useful tool to track SARS-CoV-2 and support public health policies at municipal level in Brazil. Water research, 191, p.116810.

Rački, N., Morisset, D., Gutierrez-Aguirre, I. and Ravnikar, M., 2014. One-step RT-droplet digital PCR: a breakthrough in the quantification of waterborne RNA viruses. Analytical and bioanalytical chemistry, 406(3), pp.661–667.

Randazzo, W., Cuevas-Ferrando, E., Sanjuán, R., Domingo-Calap, P. and Sánchez, G., 2020. Metropolitan wastewater analysis for COVID-19 epidemiological surveillance. International Journal of Hygiene and Environmental Health, 230, p.113621.

Sarmila, T., Sherchan, S.P. and Eiji, H., 2020. Applicability of crAssphage, pepper mild mottle virus, and tobacco mosaic virus as indicators of reduction of enteric viruses during wastewater treatment. Scientific Reports (Nature Publisher Group), 10(1).

Sherchan, S.P., Shahin, S., Ward, L.M., Tandukar, S., Aw, T.G., Schmitz, B., Ahmed, W. and Kitajima, M., 2020. First detection of SARS-CoV-2 RNA in wastewater in North America: a study in Louisiana, USA. Science of The Total Environment, 743, p.140621.

Suo, T., Liu, X., Feng, J., Guo, M., Hu, W., Guo, D., Ullah, H., Yang, Y., Zhang, Q., Wang, X. and Sajid, M., 2020. ddPCR: a more accurate tool for SARS-CoV-2 detection in low viral load specimens. Emerging microbes & infections, 9(1), pp.1259–1268.

Torii, S., Furumai, H. and Katayama, H., 2021. Applicability of polyethylene glycol precipitation followed by acid guanidinium thiocyanate-phenol-chloroform extraction for the detection of SARS-CoV-2 RNA from municipal wastewater. Science of The Total Environment, 756, p.143067.

Worley-Morse, T., Mann, M., Khunjar, W., Olabode, L. and Gonzalez, R., 2019. Evaluating the fate of bacterial indicators, viral indicators, and viruses in water resource recovery facilities. Water Environment Research, 91(9), pp.830–842.

Yang, R., Paparini, A., Monis, P. and Ryan, U., 2014. Comparison of next-generation droplet digital PCR (ddPCR) with quantitative PCR (qPCR) for enumeration of Cryptosporidium oocysts in faecal samples. International journal for parasitology, 44(14), pp.1105–1113.

